# The Mental Health Under the COVID-19 Crisis in Africa: A Systematic Review and Meta-Analysis

**DOI:** 10.1101/2021.04.19.21255755

**Authors:** Jiyao Chen, Nusrat Farah, Rebecca Kechen Dong, Richard Z. Chen, Wen Xu, Allen Yin, Bryan Z. Chen, Andrew Delios, Saylor Miller, Xue Wan, Stephen X. Zhang

## Abstract

**Objective:** In this paper, we aim to provide a systematic review and meta-analysis on the prevalence rates of mental health symptoms of anxiety, depression, and insomnia among the major populations during the COVID-19 pandemic in Africa.

**Design:** A systematic review and meta-analysis.

**Data sources:** We search and include articles using PubMed, Embase, Web of Science, PsycINFO, and medRxiv databases between Feb 202 and Feb 6th, 2021.

**Eligibility criteria and data analysis:** The meta-analysis targets the prevalence rates of mental health symptoms of major populations including frontline/general healthcare workers (HCWs), the general adult population, and medical students during the COVID-19 pandemic in Africa. To estimate the prevalence rates of anxiety, depression, and insomnia, we pooled data using random-effects meta-analyses.

**Results:** In this meta-analysis, we identify and include 28 studies and 32 independent samples from 12 countries with a total of 15,072 participants in Africa. Ethiopia (7) and Egypt (6) had the largest number of studies. While many countries including, but not limited to, Algeria, Kenya, and Ghana had a high number of COVID-19 cases, as many as three quarters of African countries have no studies. The pooled prevalence of anxiety in 27 studies was 37% (95%CI: 31-43%, *I*^*2*^ = 99.0%) and that of depression in 24 studies was 45% (95%CI: 36-51%, *I*^*2*^ = 99.5%) and that of insomnia in 9 studies was 28% (95%CI: 20-41%, *I*^*2*^ = 99.2%). The pooled prevalence rates of anxiety, depression, and insomnia in North Africa (44%, 55%, and 31%, respectively) are higher than the rates in Sub-Saharan Africa (31%, 30%, and 24%, respectively). Our analysis indicated high heterogeneity and varying prevalence rates of mental health symptoms during the COVID-19 pandemic in Africa.

**Discussion:** We discuss our findings that a) a scarcity of studies in several African countries with a high number of COVID-19 cases, b) high heterogeneity among the studies, c) the extent of prevalence of mental health symptoms in Africa to be high, and d) the pattern of mental health symptoms in Africa differs from elsewhere, i.e., more African adults suffer from depression rather than anxiety and insomnia during COVID 19 compared to adult population in other countries or regions. Hence, our findings carry crucial implications for healthcare organizations and future research to enable evidence-based medicine in Africa. Our findings also call for increased scholarly attention on Africa, the least studied continent with a limited amount of research on mental health symptoms under the COVID 19 pandemic.

**Trial registration:** CRD42020224458

## 1. INTRODUCTION

Africa, home to 1.37 billion people, is a particularly vulnerable continent due to its unique and severe limitations to handle the highly contagious COVID-19 disease^1,2^. The first confirmed case of COVID-19 in Africa appeared in Egypt on 14^th^ February 2020; and Nigeria, the most populous African country, in sub-Sahara, reported its first case on 27 ^th^ February^3,4^. As of April 11, 2021, the confirmed cases of COVID-19, disease in Africa reached to 3,139,556 with 79,404 deaths and 2.5% case fatality rate (CFR) ^5^. Africa as a whole is critically limited in terms of medical facilities and resources to deal with COVID-19^6^. Compared to other continents, Africa faces unique challenges such as lack of advanced health care facilities and intensive care units, under-staffed and over-crowded hospitals, crippling healthcare coordination and transportation, limited access to sanitary items and clean water ^7^, and the limited penetration of vaccine in its fight against the COVID-19 pandemic. The COVID-19 pandemic further threatens to disrupt the dwindling medical services and limits African peoples’ access to mental health care. Moreover, the living conditions and the high percent of uneducated adult population could increase the chances of exposure to infection and mental issues^6^. For example, a study in Ethiopia found that people using public transportation had developed and experienced general anxiety disorder^8^, In Libya a significant number of healthcare workers showed low levels of knowledge, awareness, and preparedness for COVID-19^9^. Given the situation, the prevalence of mental health symptoms in Africa during the COVID-19 pandemic is an important topic to study ^10^.

As the evidence on the mental health during the COVID-19 pandemic in Africa is accumulating, the literature has reported a heterogeneous set of findings. For example, one study found the prevalence of depression in Libya is 95.5% ^9^, and an additional study in Ethiopia found it to be 21.2% ^11^. While there are several meta-analyses on the prevalence of mental health symptoms elsewhere ^12–18^, none of the studies focus on Africa We do not know the extent of mental health symptoms in Africa, one of the least developed continent.

Hence, we aim to address this evidence gap and provide a systematic review of the literature to estimate the pooled prevalence of anxiety, depression, and insomnia in African countries during the COVID-19 pandemic. Through this systematic review and meta-analysis of studies on Africa, our work will provide a more comprehensive assessment of evidence regarding the prevalence of mental health disorders in Africa to guide and practitioners and mental health researchers during this continued global pandemic.

## 2. METHODOLOGY AND MATERIALS

### 2.1 Protocol Registration

To gain an understanding of the prevalence of mental health symptoms during COVID-19 in Africa, we conducted a systematic review and meta-analysis in accordance with the Preferred Reporting Items for Systematic Reviews and Meta-Analyses (PRISMA) statement 2019 and registered in the International Prospective Register of Systematic Reviews (PROSPERO: RD42020224458).

### 2.2 Data Sources and Search Strategy

This paper is a part of a large project on a meta-analysis of mental health symptoms during the COVID-19 pandemic globally. We performed a comprehensive literature search in the databases of *PubMed, Embase*, PsycINFO, *Web of Science*, and medRxiv from Feb 2020 to Feb 6th, 2021. We used the search query, reported in Appendix 1, and entered these keywords in each database using Boolean operators within the titles, abstracts, keywords, and subject headings (for example, MeSH terms).

### 2.3 Eligibility Criteria

In this systematic review, we included original empirical studies that have examined the impact of the COVID-19 pandemic on adults’ mental health in African countries if they met the following criteria:

a. Methodological design: cross-sectional or cohort,
b. Outcomes: reported the prevalence of depression, anxiety, or insomnia during COVID-19 pandemic,
c. Measurements/Instruments: using any validated measurement tools or scales,
d. Population: included adult participants including frontline HCWs, general HCWs, general adult population, or adult medical students from any African countries, and
e. Language: published in English.

We excluded studies based on the following criteria:

a. Population: children, adolescents, or specific niche adult populations such as patients, adults under quarantine, pregnant/postpartum women.
b. Methodological design: non-primary studies such as reviews or meta-analyses, qualitative or case studies without a validated instrument, interventional studies, interviews, or news reports.
c. Measurements/Instruments: non-validated instruments measuring mental health outcomes (i.e., self-made questionnaire) or instruments missing a validated cutoff score to calculate the prevalence rate.

A researcher (WX) contacted the authors of papers if they missed important information. For example: 1) if the surveyed population in the paper included both targeted and excluded populations in a way the prevalence rate of the relevant population cannot be identified, 2) if the prevalence rates were not reported, 3) if overall prevalence rates were reported without specifying its cutting point to determine whether it is mild above or moderate above, or 4) if the paper lacks any important information such as response rate, time-frame of data collection, sample size or gender proportions.

### 2.4 Screening and Data Extraction

One researcher (JC) exported the included articles from the databases into an EndNote library where we identified duplicates and then imported the articles to Rayyan for screening. Two researchers (BZC & AD) independently screened the imported articles based on their titles and abstracts and a third researcher (RKD) resolved any conflicts. Six coders, in pair (WX & AY, BZC & AD, RZC & SM) assessed the eligibility of each paper to be included in the review. They read the full texts to extract the relevant data based on a coding protocol into a coding book both were developed in a previous study^19^. The coding book records all the coded information such as authors’ name, year of the publication, title, publication status, sample locations, date of data collection, sample size, response rate, population, age (mean, SD, min, and max), gender proportion, instruments, cutoff scores used, and the prevalence/mean/SD of the mental health outcomes. Each paper was double-coded and crosschecked independently by a pair of coders. The pair of coders crosschecked and discussed any discrepancies. In cases where the pair of coders failed to resolve the discrepancies, a third coder (RKD) checked the paper independently to determine its coding. Another coder (RZC) double-checked important data including the population, sample size, mental health outcomes, outcome levels, instruments, and prevalence and further checked papers with unusual prevalence, cutoff scores, and numbers afterwards for sensitivity analysis.

### 2.5 Assessment of Bias Risk

We follow prior meta-analyses^20,21^ and used the Mixed Methods Appraisal Tool (MMAT) ^22^, a seven-question test to gauge the quality of the studies included in our meta-analysis. A pair of coders individually evaluated the risk of non-response bias and assessed the quality based on a representation of sample, appropriateness of measurements/instruments using the MMAT. Each article was assigned a final quality score on a range from 0 to 7. Studies with a quality score of higher than 6 were considered low bias risk, articles with a score of between 5 and 6 were classified as medium bias risk, and articles with a score of below 5 were as high bias risk. Any discrepancies regarding the quality scores were resolved either through a discussion between the pair of coders or through the lead researcher.

### 2.6 Statistical Analysis

The overall prevalence and 95% confidence intervals of mental health symptoms were pooled using Stata 16.1. Following prior literature on the prevalence of mental health symptoms, the random-effects model was used to extract the pooled estimates ^23^. We calculated the heterogeneity using the I^2^ statistic, which measures the percentage of variance resulting from the true differences in the effect sizes rather than the sampling error ^24^. We also performed subgroup analyses by the key potential sources of heterogeneity of outcomes (three types of mental health disorders), the severity of outcome (above mild/above moderate/above severe), four major population groups (frontline HCWs, general HCWs, general population, students), two regions in Africa (southern vs. Northern Africa).

## 3. RESULTS

### 3.1 Study Screening

Figure 1 illustrates the PRISMA flow chart on our search and data extraction procedures. Based on our search in the selected database and other sources, we found 6949 citations. From which, we excluded 3603 duplicates. That left us with 3346 citations. We examined the titles and abstracts of these 3346 citations and removed 2662 entries that did not meet the inclusion criteria in the screening process. Data were extracted from 684 citations based on their full text. We eliminated 505 citations in the data extraction process and coded 150 papers that included all the necessary information required to conduct meta-analyses. We also sent out two rounds of emails to the authors of 95 papers to request additional useful information required for meta-analyses and 75 responses were returned. From the authors’ responses, new prevalence data was received from 8, out of the 29 studies that lacked prevalence data. The final sample has 168 studies with sufficient information necessary to conduct a meta-analysis. Among these 168 studies, 30 articles involve participants from Africa.

**Figure 1.**
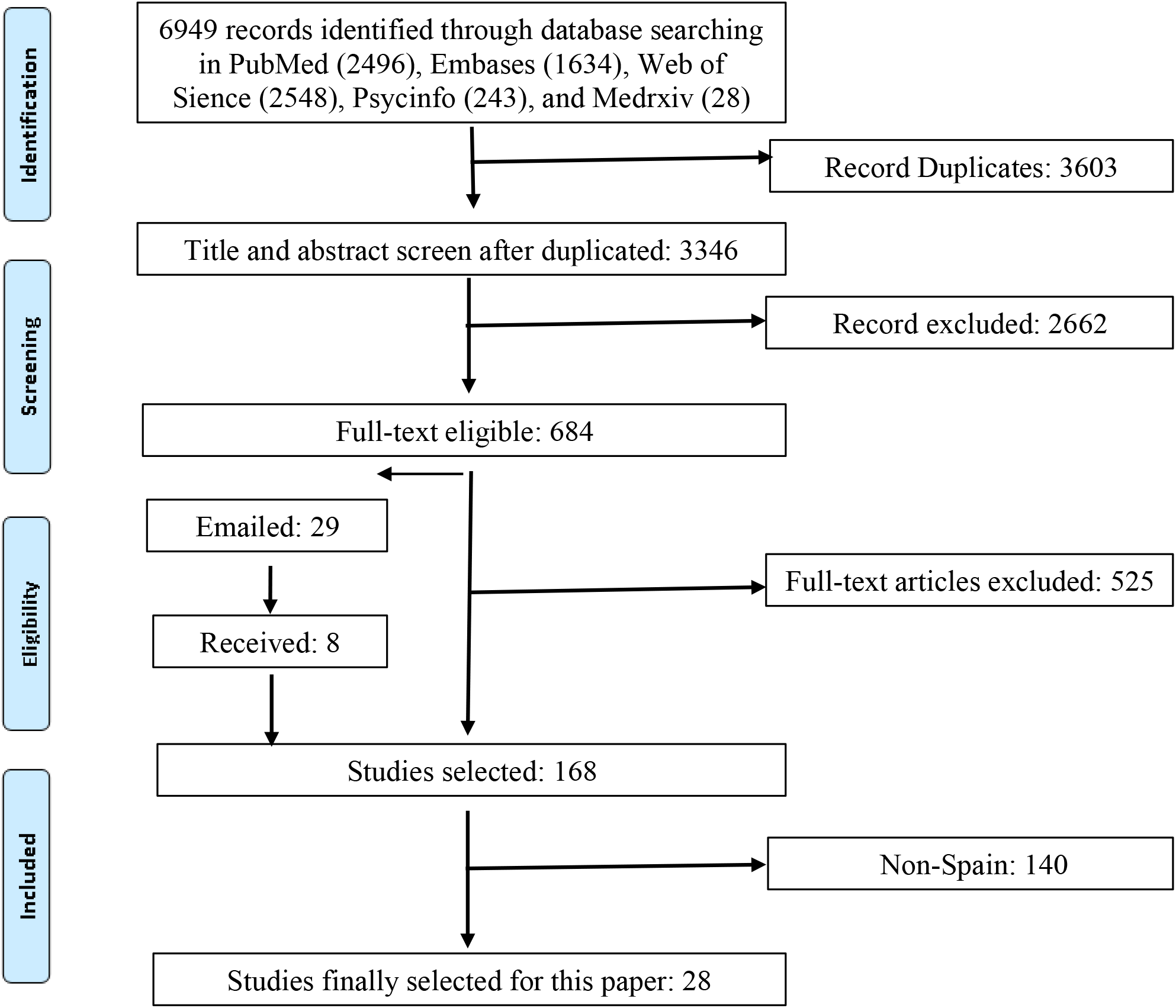
A PRISMA flow diagram.

### 3.2 Study Characteristics

In this meta-analysis, we included 28^8,9,11,25–49^ articles that contain 32 samples, 137 prevalence with a total of 15072 individual participants. Table 1 summarizes the key characteristics of the included papers. Among the 32 independent samples, 9.4% studied frontline HCWs, 37.5% studied the general HCWs, 46.9% studied the general population, and 6.3% studied medical students. 45.3% of the prevalence covered anxiety while depression is covered by 39.4% and insomnia is covered by 15.3% of the sample. Respectively, 40.3%, 32.9%, 24.1%, and 2.9% of prevalence reported at the mild above, moderate above, severe above, and overall level by the severity of the symptoms. 96.% of the studies employed cross-sectional surveys while 3.3% used cohort-based studies. All the articles were published in journals. The median number of individuals per sample was 327(range: 48 to 2,430) with a median female proportion of 56.6% (range: 0% to 100%) and a median response rate of 49.0% (range: 18.2% to 94.2%).

**Table 1.**
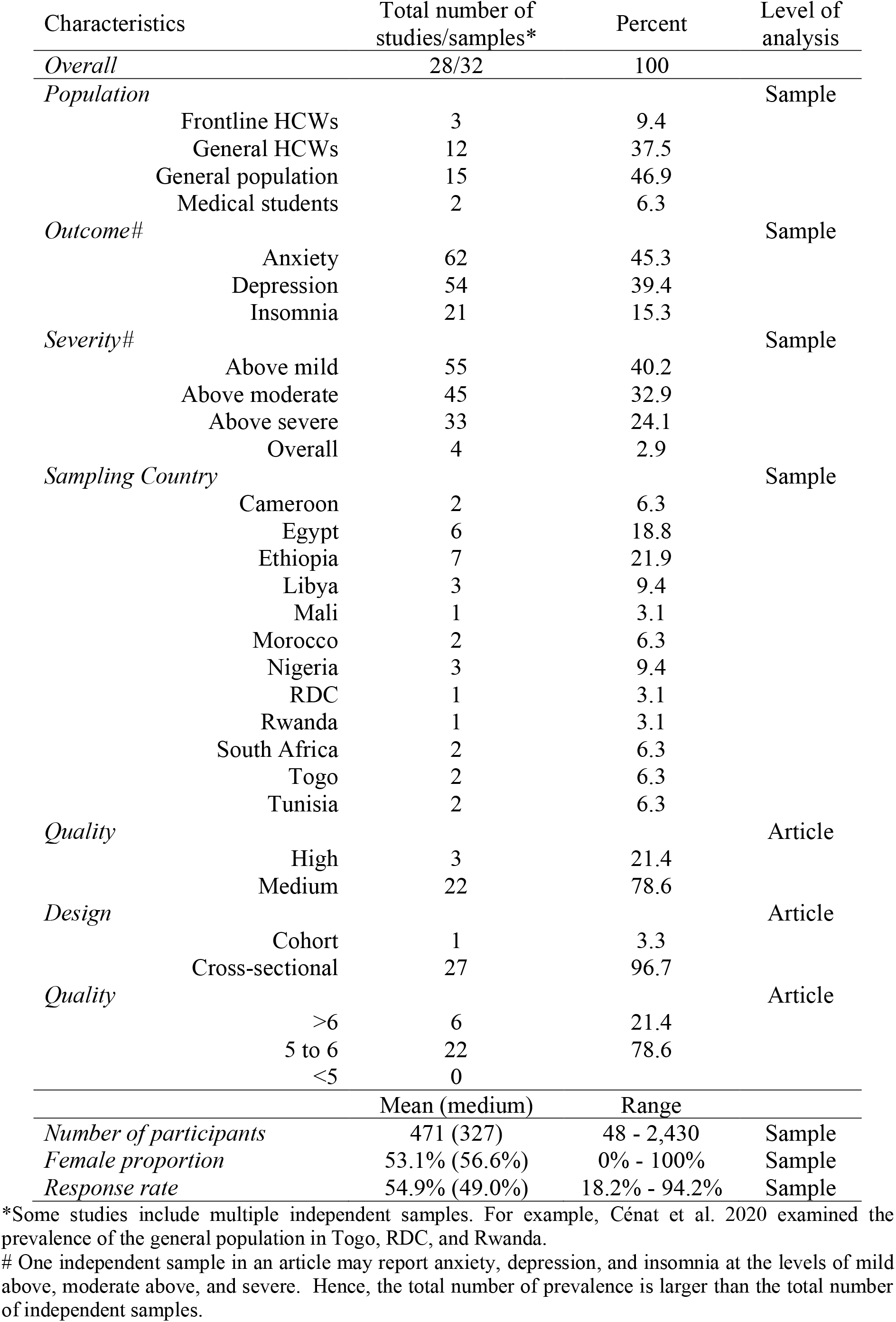
Characteristics of the studies on mental health in Africa during COVID-19 pandemic.

### 3.3 Pooled Prevalence Rates of Mental Health Disorders

The prevalence rates of the 32 samples were pooled by the subgroups (Table 2). The overall prevalence of mental health symptoms is 39% in Africa. 31 samples from 27 studies reported the prevalence of anxiety symptoms among 14,847 participants. Several anxiety instruments were used, including the Generalized Anxiety Disorder 7-items scale (GAD-7) being used the most (48.1%), followed by the Depression, Anxiety and Stress Scale - 21 Items (DASS-21) (22.2%), Hospital Anxiety and Depression Scale (HADS) (11.1%), Hamilton Anxiety Rating (HARS) (3.7%), Hopkins Symptoms Checklist (HSCL) (3.7%), Self-Reporting Questionnaire (SRQ) (3.7%), and Hospital Anxiety and Depression Scale (HADS) (3.7%). In the random-effects model, the pooled prevalence of anxiety was 37% (95% CI: 31% - 44%, I2 = 99.4%) in the 23 studies (Figure 2A).

**Table 2.**
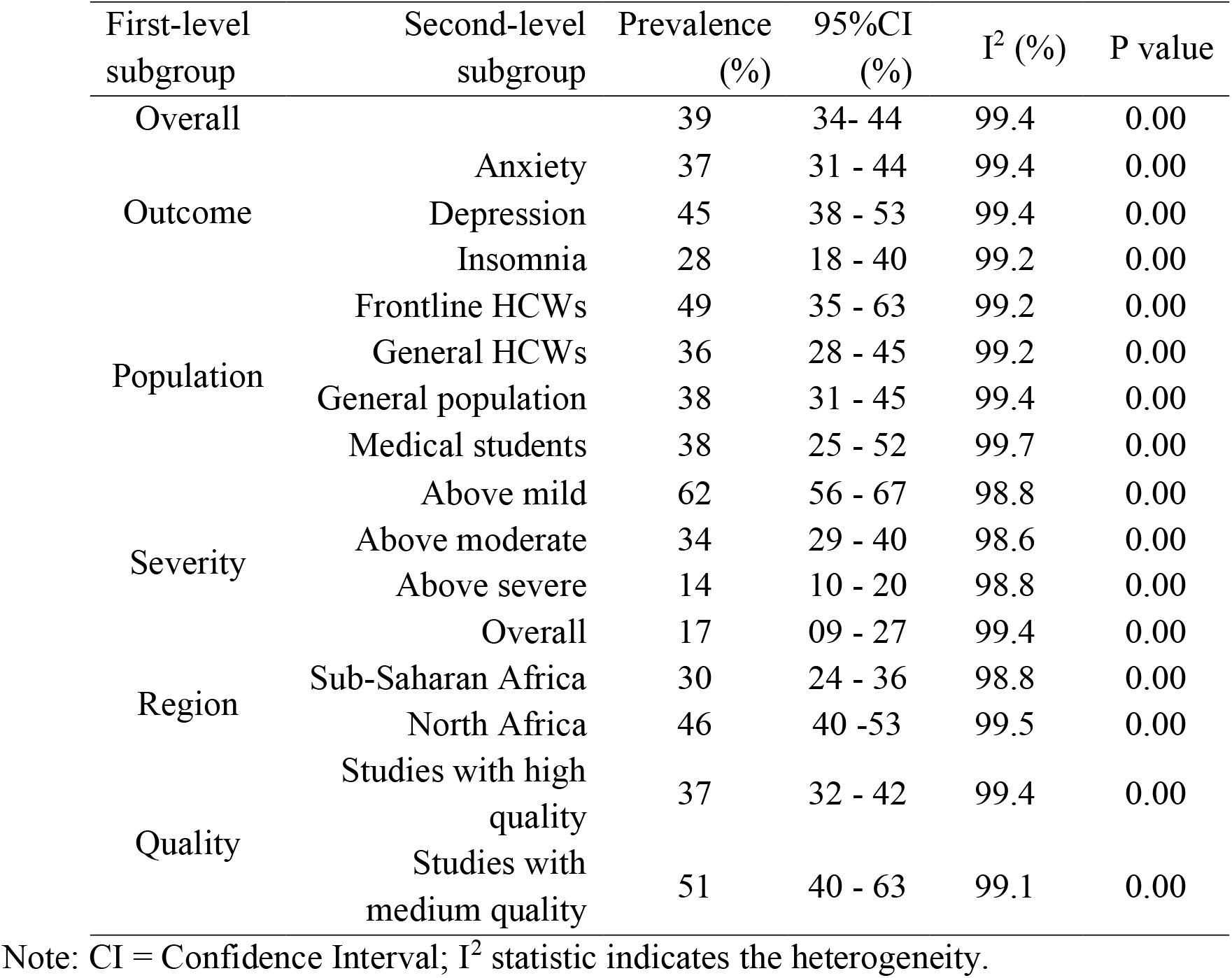
The pooled prevalence rates of mental health disorders by subgroups of population, outcome, and severity.

**Figure 2A.**
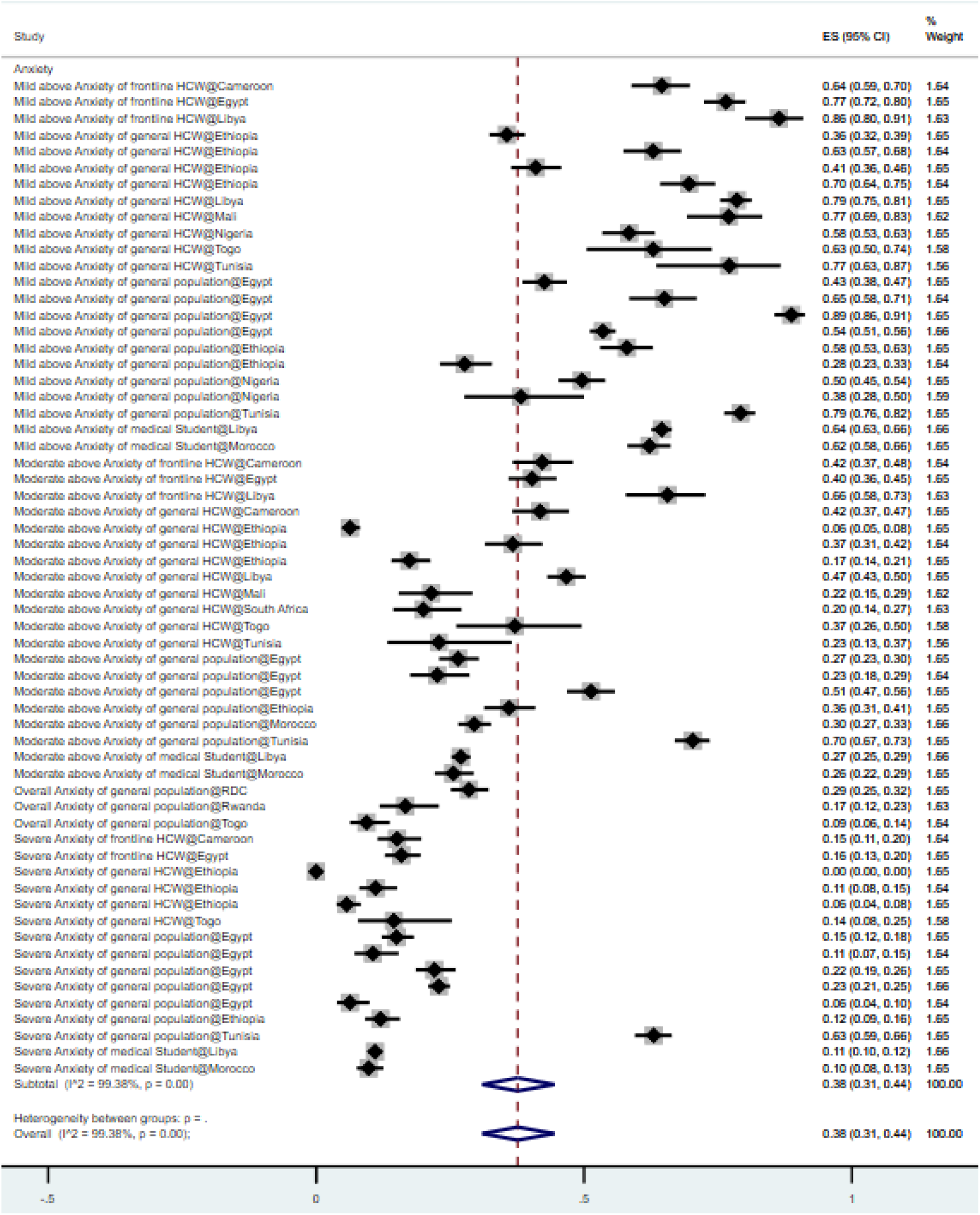
Forest plot of the prevalence of anxiety. Figure legend: The square markets indicate the prevalence of anxiety at the different level for different population. The size of the marker correlates to the inverse variance of the effect estimate and indicates the weight of the study. The diamond data market indicate s the pooled prevalence.

**Figure 2B.**
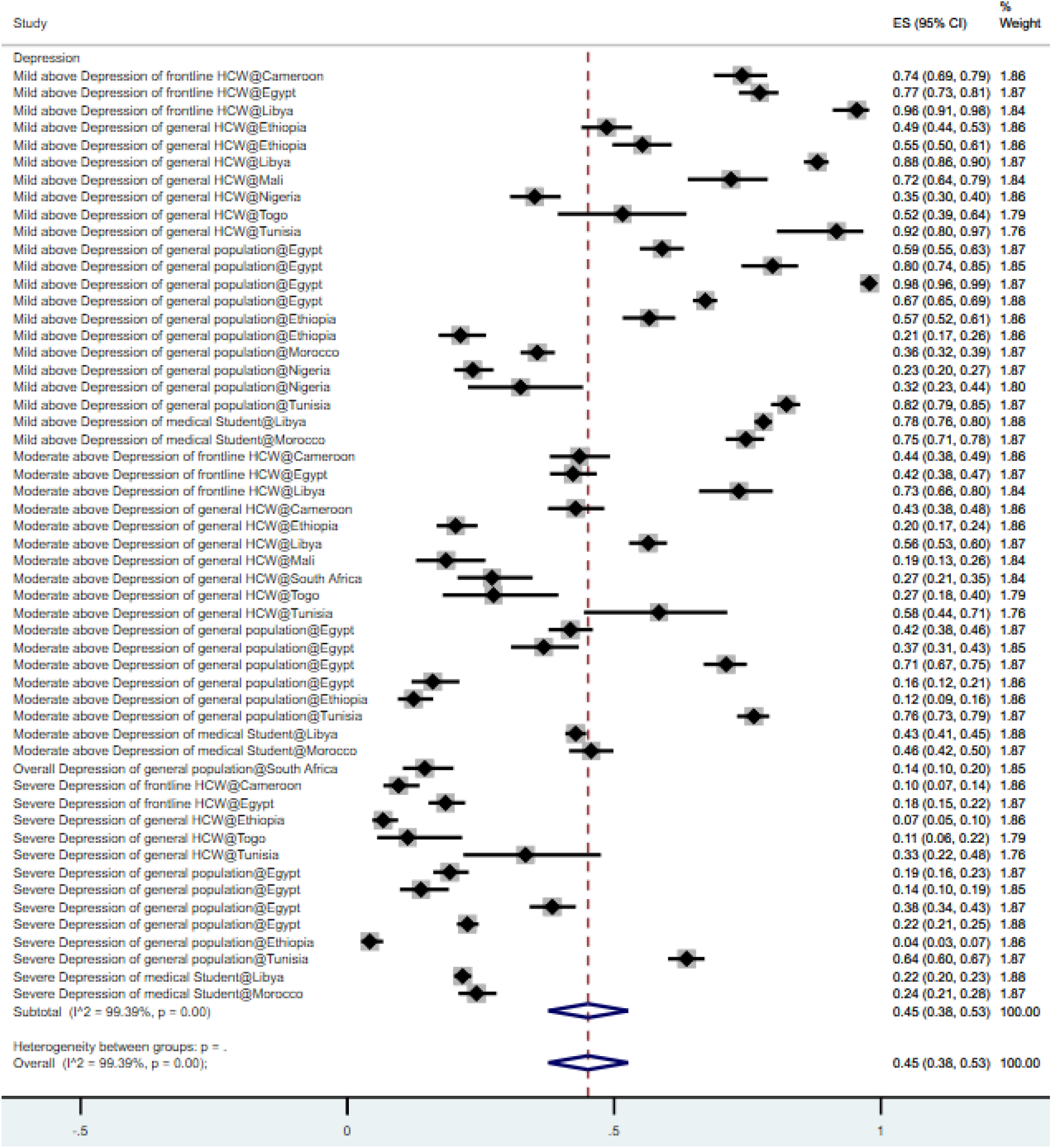
Forest plot of the prevalence of depression. Figure legend: The square markets indicate the prevalence of anxiety at the different level for different population. The size of the marker correlates to the inverse variance of the effect estimate and indicates the weight of the study. The diamond data market indicates the pooled prevalence.

**Fig 2C.**
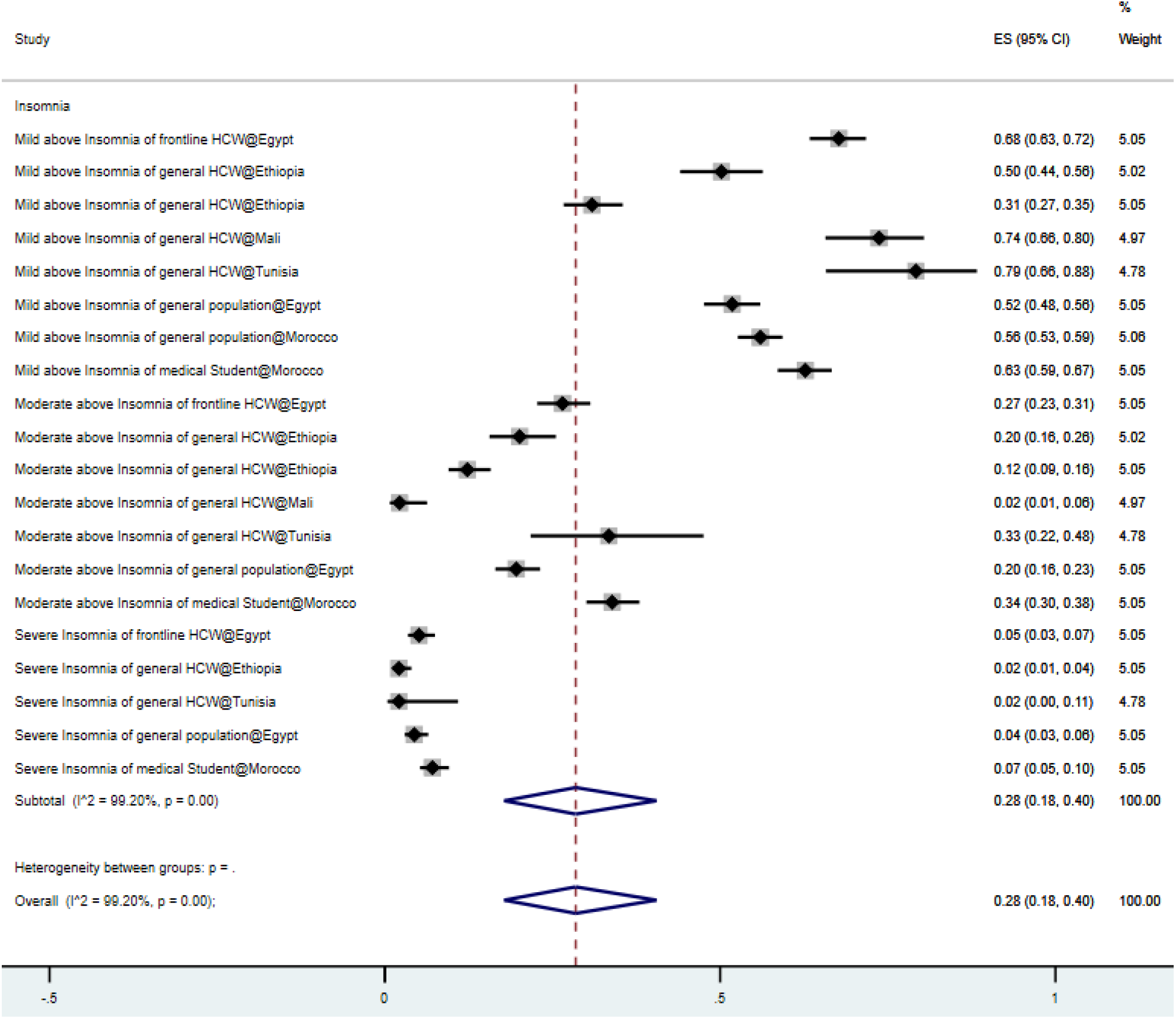
Forest plot of the prevalence of insomnia. Figure legend: The square markets indicate the prevalence of anxiety at the different level for different population. The size of the marker correlates to the inverse variance of the effect estimate and indicates the weight of the study. The diamond data market indicates the pooled prevalence.

A total of 26 samples of our total 24 articles that we reported in this meta-analysis were on depression, for a total of 12,688 respondents. Several depression instruments were used including Patient Health Questionnaire (PHQ)-9 being used the most (45.8%), followed by DASS-21 (25.0%), HADS (12.5%), Beck Depression Inventory (BDI) (4.2%), CES-D (4.2%), HADS (4.2%), and SRQ (4.2%). In the random-effects model, the pooled prevalence of depression was 45% (95% CI: 38% - 53%, I2 = 99.4%) among the 24 studies.

Ten samples of the 9 articles that we reported in this meta-analysis studied insomnia, for a total of 4144 respondents. The Insomnia Severity Index (ISI) (88.9%) and Athens Insomnia Scale (AIS) (11.1%) were used to measure insomnia. In the random-effects model, the pooled prevalence of insomnia is 28% (95% CI: 18-04%, I2 = 99.2%).

The overall prevalence rates of mental health disorders that surpassed the cutoff values of mild, moderate, and severe symptoms were 62%, 34%, and 14%, respectively. The overall prevalence of mental health disorders in frontline HCWs, general HCWs, general population, and medical students in African countries are 49%, 36%, 38%, and 38% respectively. The prevalence of mental health symptoms in North Africa is 46%, which is much higher than these of mental health symptoms in Sub-Saharan Africa (30%).

### 3.4 Study Quality

6 studies (21.4%) are of higher quality, 22 studies (78.8%) have a medium quality, and none of the studies have low quality. Subgroup analysis suggests the studies with high quality (51%) reported a significantly higher prevalence of mental health symptoms in Africa than studies with medium quality (37%) (Table 2).

### 3.5 Sensitivity Analysis

Conventional funnel plots have been found to be inaccurate for proportion study meta-analyses^38^. Due to higher sensitivity and power, a better approach for publication bias graphical representation is a DOI plot in combination with the Luis-Kanamori (LFK) index rather than using a funnel plot and Egger’s regression ^39,40^. Asymmetry is assessed quantitatively through the LFK index. Scores are ±1, between ±1 and ±2, or ±2 indicating ‘no asymmetry’, ‘minor asymmetry’, and ‘major asymmetry’ respectively. ‘No asymmetry’ is depicted in Figure 3, indicating a DOI plot and LFK index of 0.13. Therefore, the presence of publication bias is unlikely. Further sensitivity analysis was conducted on publication status and sample size; no significant influence was found.

**Figure 3.**
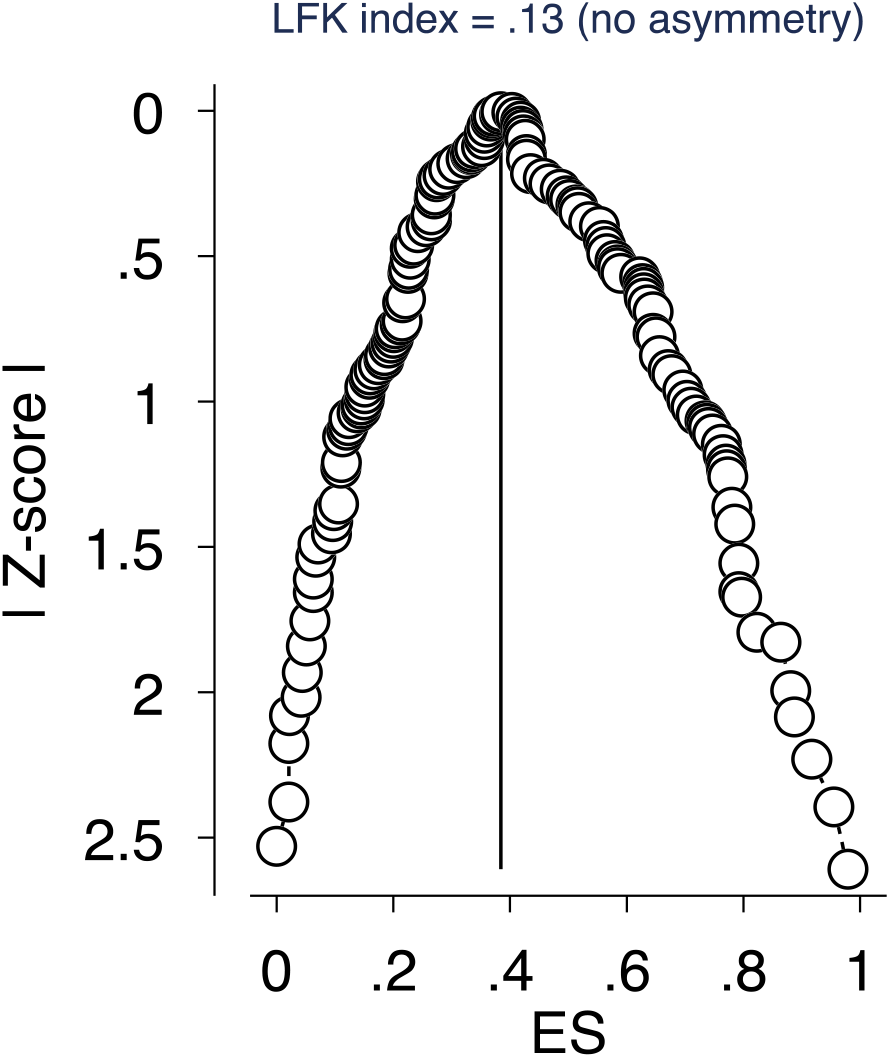
Depiction of publication bias in the baseline meta-analysis of proportion studies based on a DOI plot and the Luis Furuya–Kanamori (LFK) index-a score that is within ±1 indicates ‘no asymmetry.’

## 4. DISCUSSION

### 4.1 Comparisons of Results with Prior Meta-Analysis

Our meta-analytical findings revealed several crucial pieces of evidence on the prevalence of mental health symptoms during the COVID-19 crisis in Africa. Given this study is the first meta-analysis on the mental health symptoms during COVID-19 in Africa, it would be useful to compare our findings from Africa with some published meta-analyses in other regions. The meta-analytical evidence shows accumulatively a high prevalence of African adults suffered from depression (45%) and anxiety (37%) than insomnia (28%) – a pattern different from those in other countries. For example, more Chinese adults suffered from insomnia (19%) than anxiety (11%) and depression (13%)^19^ and more Spanish adults suffered from insomnia (52%) than anxiety (20%) and depression (23%)^50^.

The extant meta-analyses on COVID-19 mental health covered very limited regions, mostly in China and several other developed countries. Moreover, the extant meta-analytical evidence covered the literature published before May 2020 at the onset of the COVID-19 pandemic. We include articles published by February 2021 to cover more recent findings to enable better accumulative evidence on the prevalence of mental health symptoms in Africa.

Our finding on the pooled prevalence rates of anxiety in Africa mostly exceeded the reported prevalence rates by meta-analyses in other geographical areas. The pooled prevalence rate for anxiety in African population is 37%, which is significantly higher than those in China reported in Bareeqa et al. (2020) (22%; p-value <0.0001), Pappa et al. (2020) (23%; p-value <0.0001), Krishnamoorthy et al. 2020 (26%; p-value <0.0001) and Ren et al. 2020 (25%; p-value <0.0001)^12–15^, and those in Spain (20%; p-value <0.0001)^50^. However, notably, the pooled prevalence rate for anxiety in South Asian countries is significantly higher than that in Africa (41.3%; p-value <0.0001)^51^. In addition, the pooled prevalence of anxiety in Africa (37%) is higher than those in individual cross-country individual studies, such as a study of 10 countries (China, India, Japan, Iran, Iraq, Italy, Nepal, Nigeria, Spain, and the UK) (32%; p-value <0.0001)^18^ and a study in 17 countries in the regions of Asia (China, India, Japan, Pakistan, Singapore, Vietnam), Middle East (Iran, Israel), Europe (Denmark, Greece, Italy, Spain, Turkey), and Latin America (Argentina, Brazil, Chile, and Mexico) (33%; p-value <0.0001)^52^. Moreover, we find that the pooled prevalence rate of anxiety among frontline HCWs in Africa (51%) is significantly higher than Bareeqa et al. (2020) (24%; p-value <0.0001), Krishnamoorthy et al. 2020 (26%; p-value <0.0001) and Ren et al. 2020 (27%; p-value <0.0001)^12,14,15^. Similarly, we find that the pooled prevalence rate of anxiety among the general population (37%) in Africa is significantly higher than Ren et al. 2020 (24%; p-value <0.0001)^14^.

Our finding on the pooled prevalence rates of depression in Africa exceeded the reported prevalence rates by meta-analyses in most of the other geographical areas. The pooled prevalence rate for depression in African population (45%) is significantly higher than those in China reported Bareeqa et al. (2020) (27%; p-value <0.0001), Pappa et al. (2020) (23%; p-value <0.0001), Krishnamoorthy et al. 2020 (26%; p-value <0.0001) and Ren et al. 2020 (28%; p-value <0.0001)^12–15^. The pooled prevalence rate for depression in the African population (45%) is higher than those in Spain (23%; p-value <0.0001)^50^ and in South Asian countries reported by Hossain et al (2020) (34%; p-value <0.0001)^51^. The pooled prevalence for depression in Africa (45%) is also higher than the pooled prevalence in a study which had over 17 countries reported by Luo et al. (28%; p-value <0.0001)^52^ and higher than another study of 10 countries reported by Salari et al. (34%; p-value <0.0001)^18^, however, the prevalence rate for depression in Africa is lower than Italy – the country with the highest prevalence for depression (67%)^52^. Furthermore, we find that the pooled prevalence rate of depression among frontline HCW (55%) is significantly higher than Bareeqa et al. (2020) (32%; p-value <0.0001), Krishnamoorthy et al. 2020 (25%; p-value <0.0001) and Ren et al. 2020 (25%; p-value <0.0001). Moreover, the pooled prevalence rate of depression among general population (42%) in Africa is significantly higher than Krishnamoorthy et al. 2020 (24%; p-value <0.0001).

The pooled prevalence rate for insomnia in the African population (28%) is similar to those in China^19^, but significantly lower than those in Spain (52%; p-value <0.0001)^50^, Germany (39%; p-value <0.0001), Italy (55%; p-value <0.0001), and France (51%; p-value <0.0001) as well as the pooled prevalence in the 13 countries including Australia, Bahrain, Canada, Germany, Greece, Iraq, India, Mexico and USA (36% ; p-value <0.0001) as reported by Jahrami et al.^53^

The comparisons above show that the pooled prevalence rates for the African population are significantly higher than the population in most other regions, suggesting that the mental health symptoms under COVID-19 may not be uniform across regions or countries, and it is worthwhile for individual studies to understand the mental health conditions around the globe and for meta-analytical studies to pool the prevalence across regions.

The subgroup analysis that the pooled prevalence rates of anxiety, depression, and insomnia in Sub-Saharan Africa (31%, 30%, and 24%, respectively) are significantly lower than those reported in North Africa (44%, 55%, and 31%, respectively) suggests a high heterogeneous prevalence of mental health symptoms within the mega-regions of Africa. The prevalence rate differences between Sub-Saharan Africa and North Africa may arise due to lack of awareness of the danger of COVID-19 due to insufficient Covid-19 testing or the lower death rates due to the younger population in Sub-Saharan Africa ^54^. Our results call future research can examine further why such heterogeneity in the prevalence rates for the mental health symptoms occur between Sub-Saharan Africa and North Africa.

### 4.2 Practical Implications of Our Findings to Psychiatrists/Healthcare Organizations

Our meta-analysis put forward the evidence on a high proportion of mental health symptoms among the general population and healthcare workers (HCWs) under COVID-19 in Africa. Under COVID-19, the fear, worry, and uncertainty surrounding an unknown threat and containment strategies can put a burden on mental health. The higher prevalence rates of depression in the African population compared to those reported elsewhere could be due to the higher baseline prevalence rates in Africa. A meta-analysis of studies published between January 1, 2006 and July 31, 2011 showed that the pooled prevalence rate of depression symptoms and major depression in Sub-Saharan Africa is 31% and 18%, respectively ^55^. Between 2000 to 2015, 52% of the population in the African continent had some mental and substance use disorders^56^. Making issues worse, mental health symptoms received less attention in underdeveloped economies ^57^, and for instance less than 10% of people suffering from depression in low-resource settings have access to mental health treatment ^58^, despite that depression is one of the major causes of disability. The vast majority of Africa suffers from low-incomes, high mortality rates, malnutrition, high incidence of infectious diseases, and poor health facilities ^59,60^ specifically mental health resources, programs, or facilities ^57^, can present unique situations under the COVID-19 pandemic in terms of the mental health of African population.

Our findings that frontline HCWs suffered a much higher prevalence of mental health symptoms (49%) than other populations such as general HCWs (36%), general population (38%), and medical students (38%) suggest that health care organizations need to pay special attention to frontline HCWs to provide them proper mental and healthcare support during the COVID-19 crisis. Our finding that higher prevalence rates of depression (45%) than anxiety (37%) and insomnia (28%) in Africa – a pattern different from elsewhere, suggest not only the extent but also the patterns of mental health issues could be unique in Africa or at least vary across geographical regions. As such psychiatric organizations such as the World Psychiatric Association (WPA) need to consider the mental health issues in African countries differently to customize the mental health services.

### 4.3 Limitations and Future Work

Our meta-analysis is not free of limitations. First, the validity of our findings depends on the quality and reporting of the original studies included in the meta-analysis. The individual mental health studies employed a variety of instruments, cutoff scores, levels of cutoff scores to classify the severity of mental illness, and various reporting standards. For example, many studies report the overall prevalence rates without specifying which/how cutoff scores are utilized. While we have tried to reduce the additional noise and variance in the meta-analysis by paying extra attention to the severity, cutoff points, and the manners in which individual articles utilized and reported cutoff points, the profusion of different practices can lead to some biases and noise. Second, this systematic review identified empirical studies from only 12 out of the 48 Africa countries. On studies have appeared on the mental health of people in three quarters of African countries, even though those African countries are not immune to the COVID-19 pandemic. Hence, we call for research to investigate the mental health issues in Africa, the least studied continent with the majority of the countries without any studies. A possible reason is that we included articles published in English, which may result in some biases. Third, 96.7% of the studies included in this meta-analysis conducted cross-sectional surveys, whereas only 3.3% of the articles used cohort studies. We believe scholars need to focus more on cohort-based studies to examine the mental health symptoms under COVID-19 over time to further our understanding regarding how a highly contagious disease can affect psychological well-being. Finally, we only focus on studies that collected data in African countries, and we call for future meta-analyses in other countries or regions as the COVID-19 pandemic is unfolding around the globe.

### 4.4 Conclusion

This paper presents to the best of our knowledge the first systematic review, and meta-analysis on the prevalence of mental health symptoms during the COVID-19 crisis in Africa. The findings provide evidence that the prevalence of anxiety and depression is much higher in the African population compared to those reported elsewhere. Given our findings and the unique challenges Africa faces under COVID-19, we call for more studies on mental health in Africa.

## Data Availability

The data is publicly available.

## Credit author statement

**JC**: *Methodology, Validation, Formal analysis, Investigation, Data curation, Visualization, Writing – original draft, Writing – review & editing, Supervision*.

**NF**: *Writing – original draft, Writing – review & editing*.

**RKD**, *Investigation (Data)*.

**RZC**, *Investigation (Data)*.

**WX**, *Investigation (Data)*.

**AY**, *Investigation (Data)*.

**BZC**, *Investigation (Data)*.

**AD**, *Investigation (Data)*.

**SM**, *Investigation (Data)*.

**XW**: *Investigation (Data)*.

**SXZ**: *Conceptualization, Methodology, Validation, Formal analysis, Investigation, Data curation, Visualization, Writing – original draft, Writing – review & editing, Supervision*. All authors reviewed and approved the manuscript. The corresponding author attests that all listed authors meet authorship criteria and that no others meeting the criteria have been omitted.

## Transparency declaration

*The corresponding author affirms that this manuscript is an honest, accurate, and transparent account of the study being reported; that no important aspects of the study have been omitted; and that any discrepancies from the study as planned (and, if relevant, registered) have been explained*.

## Ethical approval

*Not applicable*

## Patient and public involvement

*No patient or public was involved in a systematic review and meta-analysis*

---

**Appendix 1:**
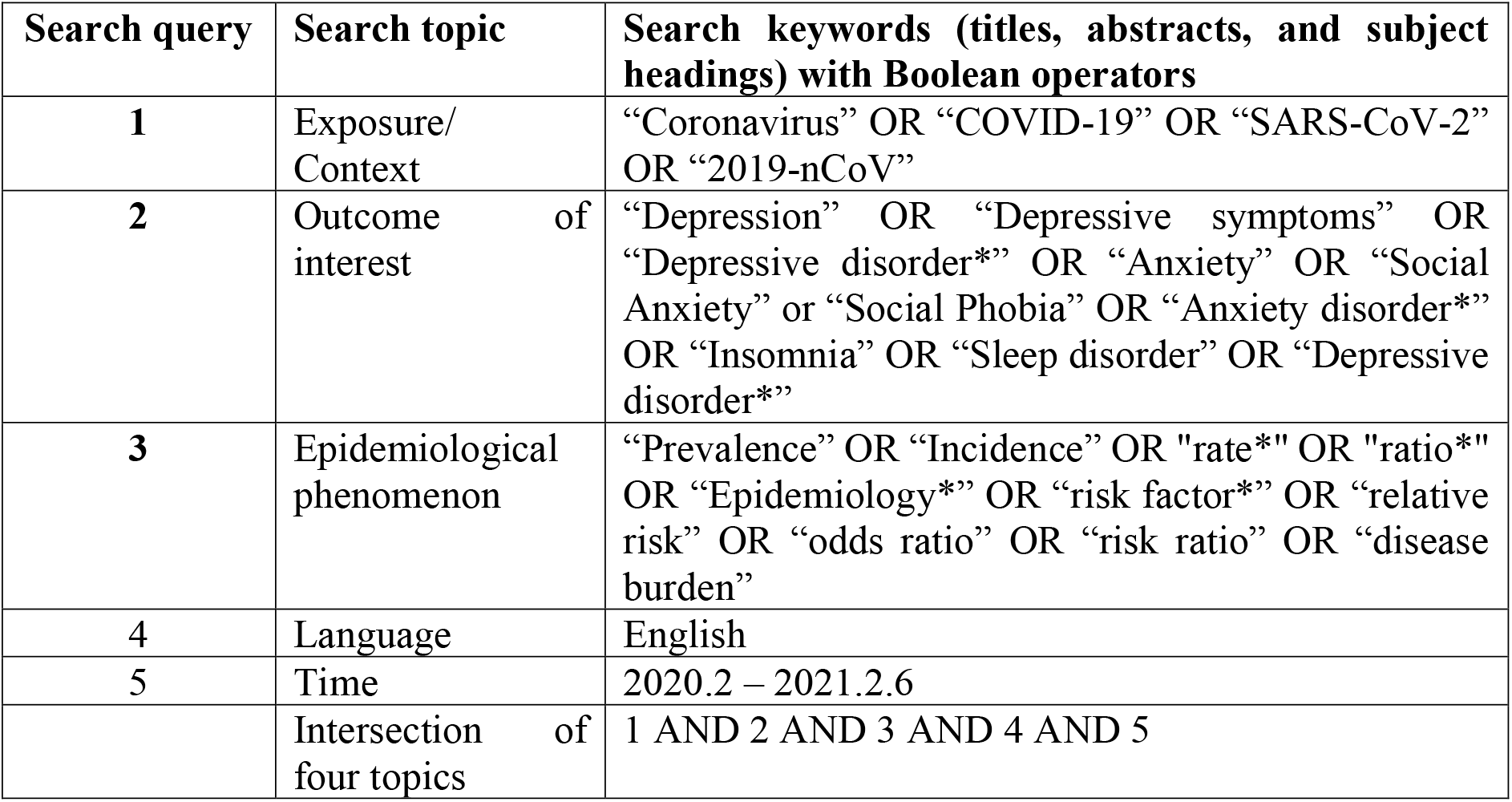
Search strategy used in this systematic review and meta-analysis.

| Search query | Search topic | Search keywords (titles, abstracts, and subject headings) with Boolean operators |
| --- | --- | --- |
| 1 | Exposure/<br>Context | “Coronavirus” OR “COVID-19” OR “SARS-CoV-2”<br>OR “2019-nCoV” |
| 2 | Outcome of<br>interest | “Depression” OR “Depressive symptoms” OR<br>“Depressive disorder*” OR “Anxiety” OR “Social<br>Anxiety” or “Social Phobia” OR “Anxiety disorder*”<br>OR “Insomnia” OR “Sleep disorder” OR “Depressive<br>disorder*” |
| 3 | Epidemiological<br>phenomenon | “Prevalence” OR “Incidence” OR "rate*" OR "ratio*"<br>OR “Epidemiology*” OR “risk factor*” OR “relative<br>risk” OR “odds ratio” OR “risk ratio” OR “disease<br>burden” |
| 4 | Language | English |
| 5 | Time | 2020.2 – 2021.2.6 |
|  | Intersection of<br>four topics | 1 AND 2 AND 3 AND 4 AND 5 |

**Appendix 2:**
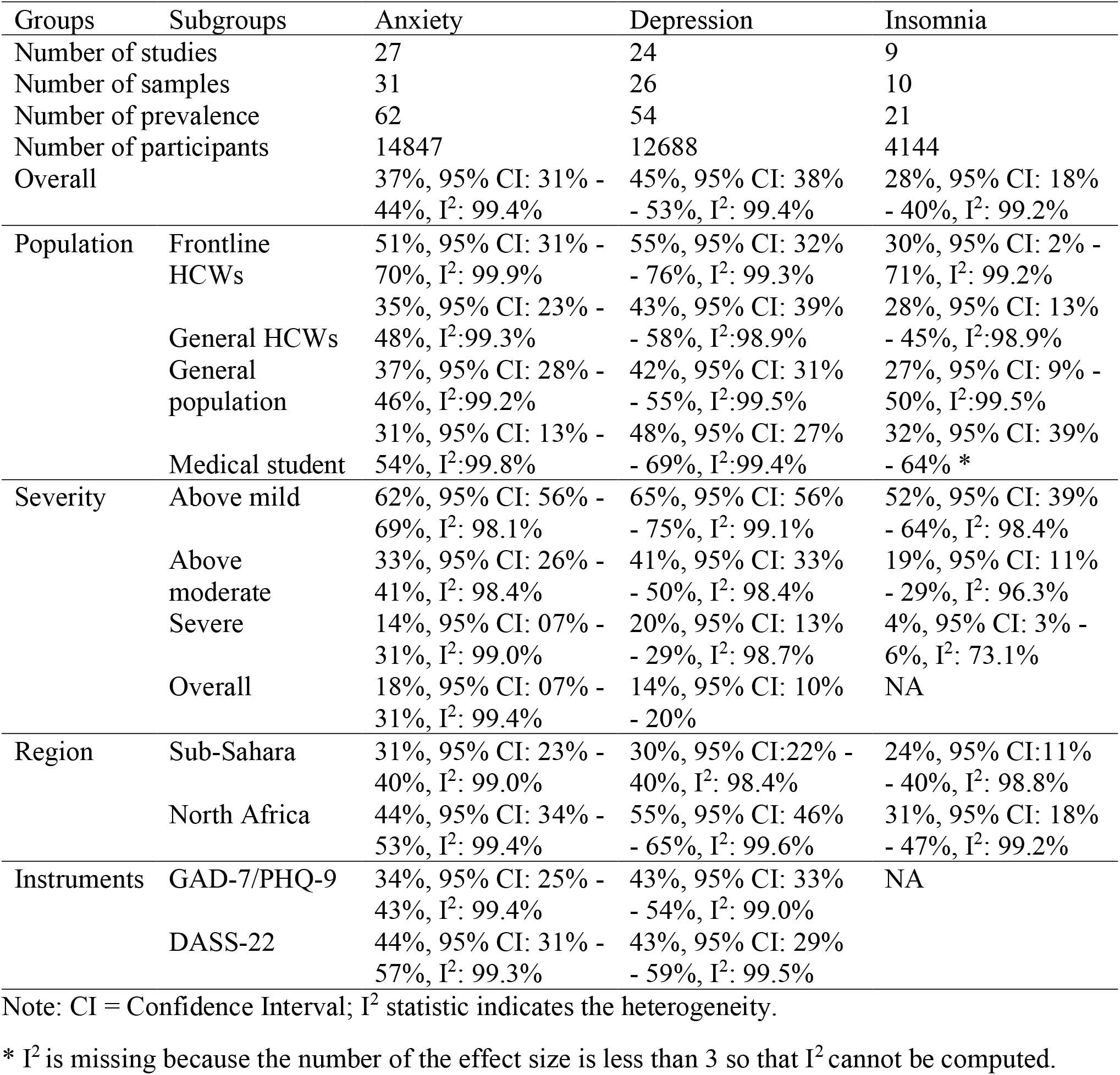
Subgroup analyses of the prevalence of anxiety, depression, and insomnia.

